# Predicting personalised risk of disability worsening in multiple sclerosis with machine learning

**DOI:** 10.1101/2022.02.03.22270364

**Authors:** Arman Eshaghi, Peter A Wijeratne, Neil P Oxtoby, Douglas L Arnold, Louis Collins, Sridar Narayanan, Charles R. G. Guttmann, Alan J Thompson, Daniel C Alexander, Frederik Barkhof, Declan Chard, Olga Ciccarelli

## Abstract

Multiple sclerosis is a heterogeneous disease with an unpredictable course. We applied machine learning to generate individualised risk scores of disability worsening and stratify patients into subgroups with different prognosis.

Clinical data and MRI scans from published randomised clinical trials in patients with relapsing-remitting and progressive MS were divided into training (n=5,483) and external validation data sets (n=2,668). We processed brain MRI scans to obtain 18 measures for lobar grey matter, deep grey matter and lesion volumes, and T1-/T2-weighted ratio of the normal-appearing white matter regions. We developed a machine learning model, called subpopulation risk stratification (SunRiSe), that combines multi-parametric clinical and MRI data to estimate individualised risk scores and stratify patients into subgroups on the basis of this risk; in particular, we entered MRI measures, the Expanded Disability Status Scale, age and gender to generate risk scores of disability worsening (i.e., the time to confirmed disability worsening). Based on SunRiSe risk scores, high-, medium-, and low-risk subpopulations were defined at study entry. We assessed whether selecting patients at high risk of disability worsening reduces sample size compared to when all risk groups were sampled together.

In both the training and external validation data sets, SunRiSe-stratified patients in three groups associated with different levels of risk of disability worsening. In the external validation data set, patients at high risk were mainly progressive MS and had more disability events compared to those at medium-risk (hazard ratio [HR]=1.34, p<0.0001) and low-risk (HR=1.51, p<0.0001). At study entry, male gender, older age, higher lesion load, higher disability, lower lobar cortical grey matter, lower normal-appearing white matter T1/T2 ratio and lower deep grey matter volumes, were the most important variables in defining the SunRiSe risk score.

The inclusion of patients predicted to be at high risk, reduced (i) duration of an event-driven trial by an average of 4.5 months (±2.1 months); (ii) the number of participants in a randomised trial by approximately 200, with 80% statistical power to detect a 30% treatment effect.

Machine learning provides a personalised risk score that can identify patients who have the greatest risk of disability worsening and therefore should be treated with the most effective medications and monitored more closely. Risk stratification allows the enrichment of clinical trials with patients more likely to worsen, and thereby reduces trial duration and sample size.

## Introduction

Predicting the disease course at an individual level in clinical practice is essential to achieve precision medicine. Predicting the future disease course in clinical trials allows recruiting individuals with similar characteristics and reducing participants needed to achieve statistical power by (1) increasing the chance of reaching the end point and (2) reducing between subject variability. Machine learning tools hold great promise to predict disease course in the clinical and research setting, because they can integrate routinely available information from different sources to objectively predict the future course of diseases at an individual level^1–6^.

Multiple sclerosis (MS) is the most common demyelinating neurodegenerative disorder in young adults in Europe and the USA^7^. Previous studies have used clinical and imaging measures, alone or in combination, to predict short- and long-term MS disability^7–10^. These studies have used group-level statistical approaches, but more recent machine learning methods are more adept at predicting prognosis at the individual level^8, 11–16^. Despite these advances, robust validations in external cohorts and clinical trials are lacking. It remains unclear whether we can generate meaningful risk scores for individuals to assist in decision making in clinical practice at an individual level and recruit patients at similar risk of disease worsening in clinical trials to maximise treatment effects or reduce adverse events. In our recent work^17^, we used a cross-sectional method to differentiate data-driven subtypes of MS on the basis of patterns of MRI changes. Our previous model based on Subtype and Stage Inference (SuStaIn) algorithm ^18^ could not include clinical measures that fluctuate over time (non-monotonic variables). Here, we aimed to expand our previous cross-sectional MRI-only models with longitudinal and clinical information and learn the relationship between baseline MRI and clinical measures with the time to disability worsening.

Machine learning integrates information from multiple sources, such as brain MRI scans and clinical measures, to enable objective prediction of future clinical course. Survival modelling estimates the effects variables have on the time to reach a pre-defined event. It is currently applied at the group level to infer the average risk of reaching specific clinical events. Recent advances in machine learning have enabled personalised predictions and provided individual risk scores^3^. These models hold great promise to provide prognostic information in advance of recruitment, and in future, may help in designing and recruiting participants in machine learning-assisted clinical trials.

We introduce a new model called SunRiSe (subpopulation risk stratification inference) to generate an individual risk score from a combination of quantitative MRI and clinical variables at study entry and predict the chance of developing disability worsening over time. Our experiments test the hypotheses that: (1) calculating SunRiSe risk scores with quantitative MRI and clinical data at study entry, improves prediction of disability outcomes when compared with the use of clinical data alone, and (2) screening in patients with a predicted high-risk for clinical worsening increases the statistical power of clinical trials to detect treatment effects.

## Methods

### Overview

We introduce a machine learning model called SunRiSe based on MRI measures of regional neuroanatomical volumes, T1/T2 in the normal-appearing white matter regions, lesion load and clinical information at study entry. This model generates risk scores for disability worsening at an individual level. We applied SunRiSe to unseen trial cohorts to confirm the prognostic ability of the model. We then used these risk scores to select higher-risk patients in simulated clinical trials and determined how SunRiSe can accelerate clinical trials by reducing the sample size. Below, we will explain the study setting, SunRiSe model development and related experiments.

### Data: study design, participants, and data sources

This was a retrospective study on clinical trials collected under the auspices of the International Progressive MS Alliance (https://www.progressivemsalliance.org/). We used clinical and MRI data from 17 studies which included MS randomised-controlled trials (RCTs) of primary progressive MS (PPMS)^19–23^, secondary progressive MS (SPMS)^24–29^, and relapsing-remitting MS (RRMS)^20, 30, 31^, and observational cohorts with mixed MS subtypes, which were previously analysed and published^17^. We split our data set *a priori* into 14 studies for training and cross-validating our model, and 3 RCTs to assess the model validity and generalisability. The training data sets consisted of 11 clinical trials and three observational cohorts. The external validation data set consisted of three clinical trials: one in PPMS^19^, one in SPMS^24^ and one in RRMS^32^.

### Ethical approval and patient consent

Each RCT and observational study had received ethical approval, and participants had given written informed consent at the time of data acquisition. In addition, the Institutional Review Board at the Montreal Neurological Institute (MNI), Quebec, Canada, approved this study (Reference number: IRB00010120) under the auspices of the International Progressive MS Alliance.

### Brain MRI Protocol and image processing

We used the following brain MRI sequences available in each trial cohort: T1-weighted, T2-weighted, and Fluid Attenuated Inversion Recovery (T2-FLAIR) MRI. We used brain 2D and 3D T1-weighted scans to segment grey and white matter tissues, T2-FLAIR to segment lesions, and T2-weighted scans, together with T1-weighted scans, to obtain T1/T2 ratio. Details of MRI protocols are explained in publications associated with each dataset^19–22, 24–28, 30–37^ and our previous work^17^.

We used regional brain volumes, lesion volume, and T1/T2 ratio values of the normal-appearing white matter (NAWM), processed and quality checked and explained in detail in our previous work^17^. The image processing included correction for scanner inhomogeneity with N4 Bias Field Correction^38^ (part of Advanced Normalisation Tools or ANTs^39^). We used a fully automatic T2-FLAIR lesion segmentation^40^ tool and filled hypointense lesions with normal-appearing tissue in T1-weighted MRI to reduce segmentation errors with the NiftySeg software^41^, and segmentation of brain tissue into GM, WM and CSF with the Geodesic Information Flows (GIF) software version 3.0^42^. We extracted the following variables based on brain regions:

- Volumes of the frontal, parietal, temporal, and occipital grey matter, limbic cortex, cerebellar grey matter and white matter, brainstem, deep grey matter and cerebral white matter. To obtain lobar GM volumes, we merged GM regions according to the brainCOLOR protocol (https://mindboggle.info/braincolor/).
- Total T2 lesion volume
- T1/T2 ratio (a proxy for the white matter integrity) of the NAWM in the corpus callosum, frontal, temporal, parietal, occipital lobes, cingulate bundle and cerebellum.

### Defining the event outcome

The Expanded Disability Status Scale (EDSS)^43^, which is a measure of neurological disability, was scored according to the individual study protocol. We used the EDSS at least one month after a protocol-defined relapse. We defined ‘confirmed disability worsening’ (CDW) as a worsening of EDSS sustained on subsequent visits for at least 12 weeks. We defined EDSS worsening as: ≥1.5-point increase from a baseline EDSS of 0, ≥1-point increase from a baseline score of 0.5 to 5.5, and ≥0.5-point increase from a baseline score >5.5.

### Preparing variables

We adjusted MRI measures for clinical and demographic variables to disentangle the effects of demographic and clinical variables on MRI measures and report the independent contribution of variables to model predictions. We constructed univariate regression models in which age, age squared, gender, and the EDSS at study entry were the independent variables and each MRI measure was the dependent variable. Disease duration was not included in these models because it was highly correlated with age. We extracted the residuals of MRI measures and used them for further analysis to ensure that MRI measures were contributing independently of demographic and clinical outcomes in predictive models (see below).

### Model development

#### Developing and training SunRiSe

SunRiSe is a consensus of three diverse models that combine into a single model to provide superior performance when compared to each individual model, as shown in Figure 1. Combining different models to improve performance or ensemble learning is well-established and has previously been applied to other neurodegenerative disorders^2^ and MS^44^. SunRiSe is trained using longitudinal data, but requires only cross-sectional data to perform predictions after training. The novelty of SunRiSe is using neural network and classical mixture models to balance the accuracy of predictions with generalisability. The three components were:

1. A deep neural network survival model with the “DeepHit” architecture (Lee et al.^3^)
2. Trajectory-based classifier with multilevel mixture models^45^
3. Gradient boosting survival trees

**Figure 1.**
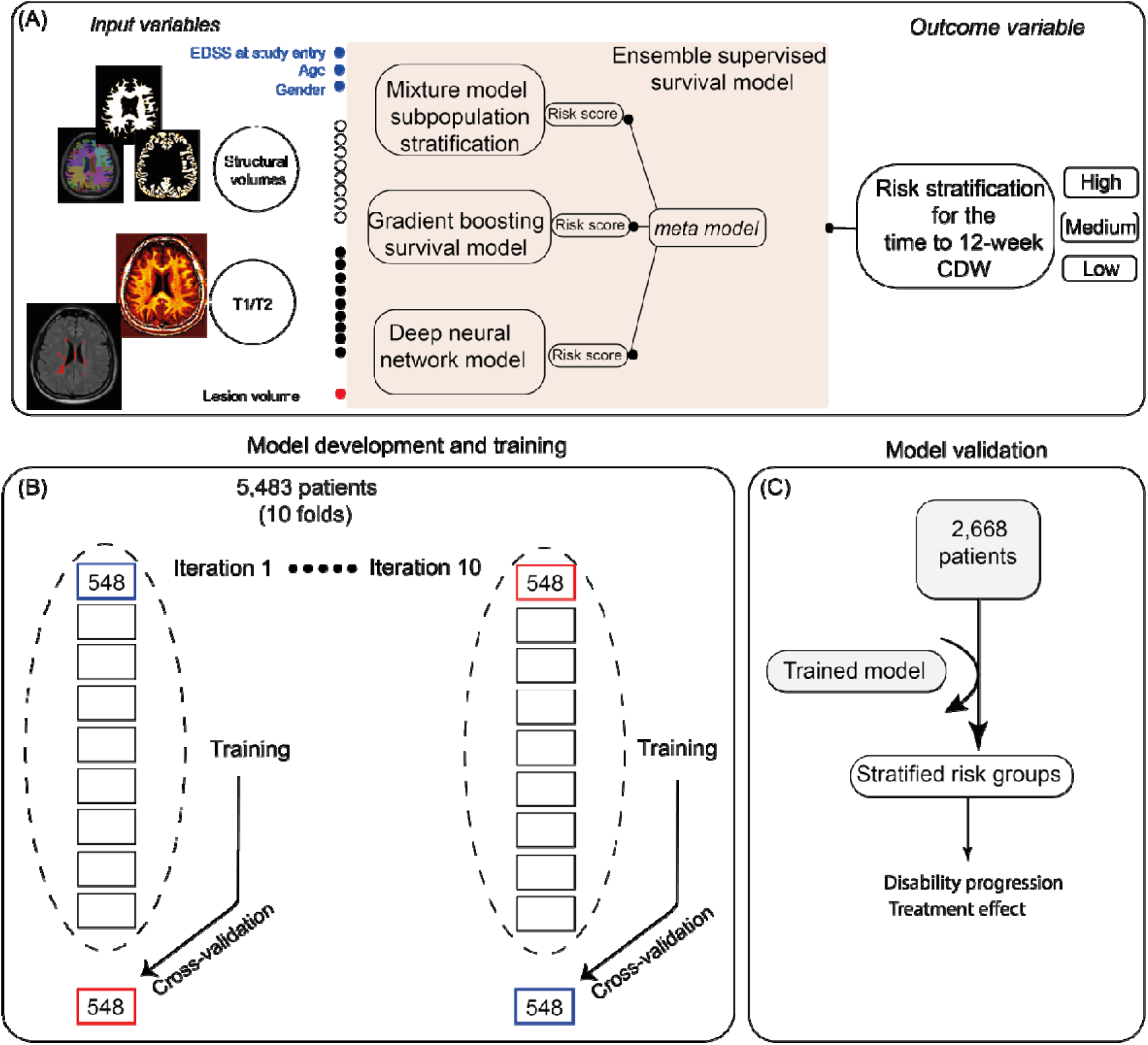
Overview of the training and validation. This flowchart shows how we trained the ensemble SunRiSe model. (A) shows the input variables from different modalities (each circle represents a variable and different colours represent modalities). Each model generates a risk score, the meta-model generates a final risk score from the three risk scores. The risk scores are grouped into high, medium and low risk. The threshold to define these three groups are based on the tertiles of the risk scores in the training data set; (B) shows the training and cross-validation of the model; (C) shows how we applied the trained model to the data at study entry in the external validation trial cohorts to assess validity and generalisability of the risk stratification framework. *Abbreviations*: CDW, confirmed disability worsening.

DeepHit is a deep neural network with an architecture designed for survival modelling. It captures non-linear relationships across predictors better than Cox regression models^3^. DeepHit is explained in the Supplemental Material and in more detail elsewhere^3^. The second model was a longitudinal (multilevel) mixture model. The mixture model is a probabilistic model that explores the presence of subpopulations without the need to rely on arbitrary thresholds (unsupervised learning). We used this model to answer the question: how many subgroups of EDSS worsening can best explain its pattern of variability over time? We explored the optimal number of subgroups with a similar pattern of EDSS worsening in the training data set, trained a classifier on baseline variables and used the probability of belonging to subgroups to weight predictions of a Cox survival model in external validation data. Time was nested as a random effect variable within the ‘subject’ variable to adjust for repeated measures in multilevel mixture models^45, 46^. EDSS at each visit was the outcome in the mixture model, while time was the fixed effect variable. Our justification for using mixture models in addition to the neural network model was that mixture models are less sensitive to data heterogeneity and thereby will increase the generalisability of our models to external, unseen data. In the third model, we used a gradient boosting approach with Cox proportional hazards models because they are among the most successful tools for classification and survival modelling^47^. The Supplemental Materials explain the methodological details of these models.

We developed our models with two sets of variables: one with 18 MRI variables (called MRI-only model henceforth) and another with 21 variables, which included the previously mentioned 18 MRI variables, EDSS, gender, and age at study entry (see Figure 1). The outcomes were: (1) the time to the beginning of 12-week CDW for those who experienced CDW; (2) the time present in the study for those who did not experience CDW (i.e., right-censored); and (3) a categorical variable, which indicated whether the patient experienced CDW or not (i.e., the patient completed or dropped out of the study without experiencing CDW [being censored]).

### Classification of patients into risk groups

We ranked the predicted risk scores for developing CDW at two years from each of the three models and calculated one “consensus” risk score for each patient (referred to as the SunRiSe risk score henceforth) using a linear model. We chose two years because it is a common duration for phase 2 and 3 clinical trials. We used a ‘meta’ survival model to predict the disability outcomes from individual models’ risk scores. To avoid *post hoc* and arbitrary thresholds in defining subgroups, we used multilevel mixture models which showed the three subgroups that optimally described EDSS worsening (see Supplemental Methods). We defined thresholds of high-, medium- and low-risk groups based on tertiles of the consensus generated risk scores in training data set (to avoid circular reasoning we did not use external validation data set) and defined these three risk groups in the external validation set. We used the Cox-regression models to compare the risk of developing CDW over time across these three groups, separately in training and external validation data sets. We used Kaplan-Meier curves to visualise the relationship between CDW events and predicted risk groups. We also looked at the relationship between the SunRiSe risk scores with EDSS at study entry and over time. For this analysis, we used a mixed-effects model in which EDSS was the outcome, with risk score and time (and their interaction) as predictors. The “subject” was the random effect variable used for nesting observations in the mixed effects model.

### Treatment response calculation

From the three clinical trials in the external validation data set, we used the ASCEND and ORATORIO clinical trials to calculate the treatment response and set BRAVO aside. Our justification for this was that the BRAVO clinical trial was a negative study^32^, while ORATORIO was a positive trial and ASCEND had evidence of treatment response in secondary outcome measures.^19, 24^ For each subgroup (high-risk, medium-risk and low-risk), we used mixed-effects models in which the outcomes were EDSS, Timed-Walk Test and 9-Hole Peg Test. Time and RCT arms (placebo vs treatment) and interaction between time and RCT arm were predictors. We used the beta coefficient of the interaction term (Time x RCT arm) as the treatment response.

### Selecting high-risk patients to enrich event-driven clinical trials

We determined whether selecting patients at high risk of developing CDW could increase statistical power compared to clinical trials without risk stratification (i.e. we sampled high, medium and low risk groups with equal weighting). We randomly sampled patients’ first visits in the placebo groups of the three clinical trials in the external validation data set and calculated the power to detect 12-week CDW. For this, we used the trained SunRiSe model to predict risk scores and sampled 1,300 patients based on model-predicted risk scores. We performed two simulations for event-driven and clinical trials with pre-defined sample sizes:

1. Event-driven clinical trials: We sampled patients classified as high-risk from the placebo groups. In both scenarios, we used 1, 2, and 3 years of follow-up to detect CDW with a simulated hazard ratio of 0.7 (approximately 30% treatment effect in line with phase 3 clinical trials showing treatment effects in progressive MS^19, 48^) and reported the statistical power at a two-sided alpha level of 0.05. We also calculated the power for clinical trials when patients were sampled randomly. We used a constant screening time of 12 months for all the sample size calculations.
2. Full-duration (“regular”) clinical trials: We repeated the simulations to calculate the number of participants needed to achieve 80% statistical power over three years of follow up with the same simulated treatment effects.

We used Schoenfeld’s approximation method^49^ and repeated the sampling with replacement for 5,000 times to obtain standard deviations (i.e. bootstrapping^50^).

### Software toolboxes

We used Pytorch version 1.5.0, PyCox version 0.2.2, and Sksurv version 0.15.1 for machine learning experiments^51^. We used MPlus version 8.4 for longitudinal mixture modelling^45^. We used R Version 4.0.3 for all the remaining statistical analyses and plotting of results^52^ and gsDesign package version 3.2.1 for the power analysis.

### Data Availability

The data sets are controlled by pharmaceutical companies. Requests to access data can be forwarded to data controllers listed in our previous publication^17^. Processed CSV files can be from the corresponding author by any qualified investigator for reproducing the results of this study.

## Results

### Demographic characteristics

There were 5,483 patients with RRMS (3,194), SPMS (1,182) and PPMS (1,107) in the training data set. There were 2,668 patients in the external validation data set including1,116 RRMS, 851 SPMS, and 701 PPMS. The average age (±standard deviation or SD) for RRMS was 36.9 years in the training data set and 37.60 in the external validation data set, 53.07 (±7.40) for SPMS in the training set and 46.67 (±7.67) in the external validation set. The average age was 49.26 (±8.47) years for PPMS in the training set and 44.57 (±8.00) in the external validation data set. The training and external validation data sets did not differ in age, EDSS, gender distribution, and disease duration (p-values>0.05 for all comparisons). **Table 1** shows the demographic and clinical characteristics of patients at study entry.

**Table 1.**
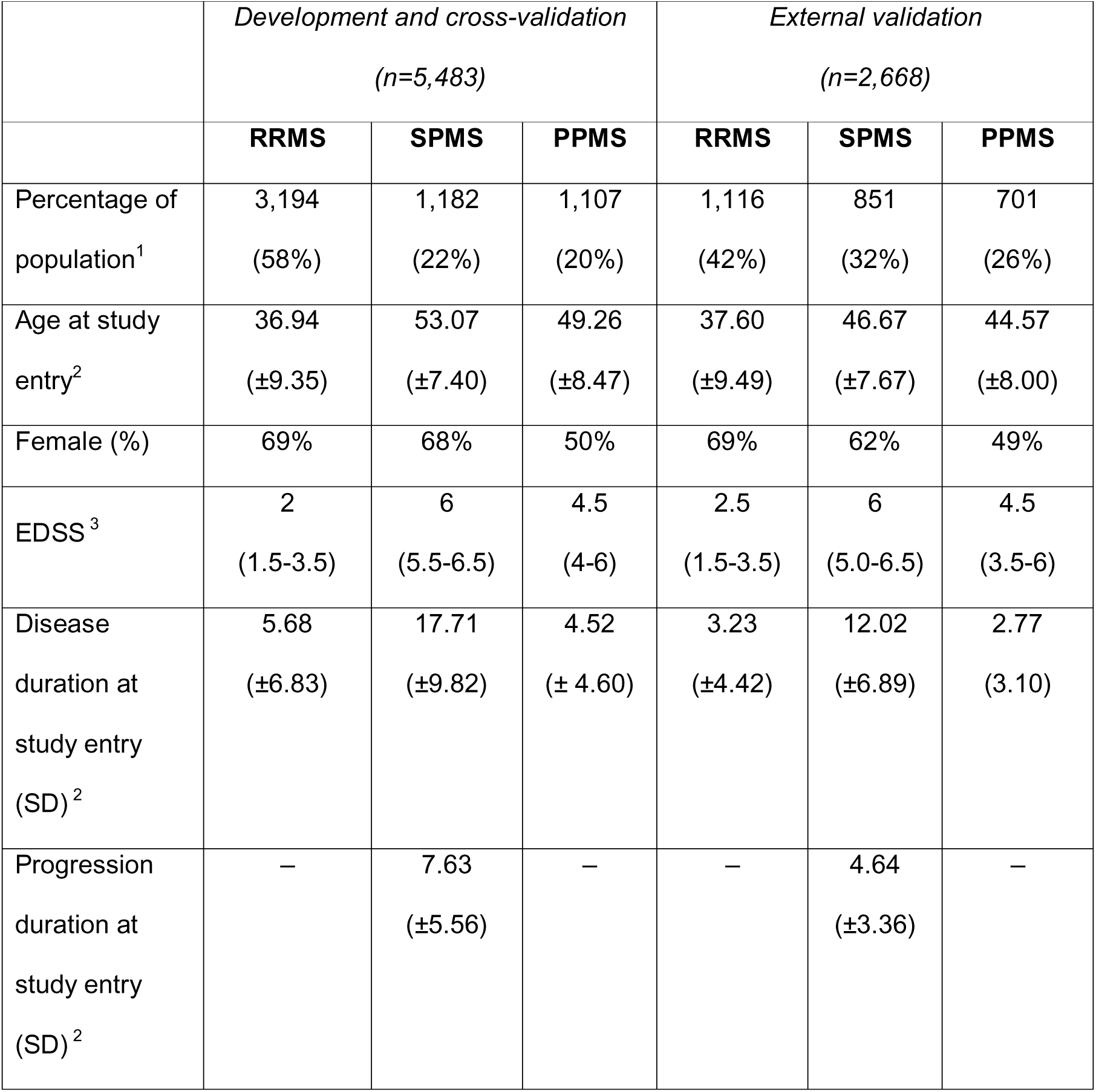
Patient characteristics of the development and external validation data sets.

|  | *Development and cross-validation*<br>(n=5,483) |  |  | *External validation*<br>(n=2,668) |  |  |
| --- | --- | --- | --- | --- | --- | --- |
|  | **RRMS** | **SPMS** | **PPMS** | **RRMS** | **SPMS** | **PPMS** |
| Percentage of population<sup>1</sup> | 3,194<br>(58%) | 1,182<br>(22%) | 1,107<br>(20%) | 1,116<br>(42%) | 851<br>(32%) | 701<br>(26%) |
| Age at study entry<sup>2</sup> | 36.94<br>(±9.35) | 53.07<br>(±7.40) | 49.26<br>(±8.47) | 37.60<br>(±9.49) | 46.67<br>(±7.67) | 44.57<br>(±8.00) |
| Female (%) | 69% | 68% | 50% | 69% | 62% | 49% |
| EDSS<sup>3</sup> | 2<br>(1.5-3.5) | 6<br>(5.5-6.5) | 4.5<br>(4-6) | 2.5<br>(1.5-3.5) | 6<br>(5.0-6.5) | 4.5<br>(3.5-6) |
| Disease duration at study entry (SD)<sup>2</sup> | 5.68<br>(±6.83) | 17.71<br>(±9.82) | 4.52<br>(± 4.60) | 3.23<br>(±4.42) | 12.02<br>(±6.89) | 2.77<br>(3.10) |
| Progression duration at study entry (SD)<sup>2</sup> | – | 7.63<br>(±5.56) | – | – | 4.64<br>(±3.36) | – |

#### Predicting the risk of CDW: added value of MRI with multimodal prediction models

*Using clinical data at study entry,* in the training data set, patients classified as high-risk were more likely to experience 12-week CDW compared to the low-risk group (HR = 1.27, 95% CI =1.1 to 1.44, p < 0.001). Patients classified as medium-risk were more likely to experience CDW compared to the low-risk group (HR=1.14, 95% CI=1.02 to 1.55, p<0.03). Similarly, in the external validation data set, patients predicted to be high-risk were more likely to experience CDW than those classified as low-risk (HR=1.31, 95% CI = 1.07 to 1.61, p=0.007). This means that patients classified by the model as high-risk were approximately 31% more likely to experience CDW than the low-risk group. On average, patients predicted to be high-risk experience more CDW events than the medium-risk group (HR=1.16, 95% CI: 0.94 to 1.43), but this was not statistically significant (p=0.15).

### Using MRI at study entry

#### Predicting risk using only MRI at study entry in training and external validation data sets

In the training data set patients classified as high-risk were more likely to experience 12-week CDW compared to the low-risk group (HR = 1.69, 95% CI =1.48 to 1.92, p < 0.001) and more likely to experience CDW compared to the medium-risk group (HR = 1.52, 95% CI =1.34 to 1.72, p<0.001). This means that patients classified by the model as high-risk were more likely to develop CDW, by approximately 52% relative to the medium-risk group and 69% relative to the low-risk group. There was no significant difference in reaching the 12-week CDW between the medium and low-risk groups, although this risk indicated on average greater CDW events in patients classified as medium risk (HR= 1.10, 95% CI = 0.96 to 1.24, p=0.13). Similarly, in the external validation data set, patients classified as high-risk were more likely to experience EDSS progression than those classified as low-risk (HR=1.21, 95% CI = 1.00 to 1.48, p=0.05). This means that patients predicted to be at high-risk were approximately 21% more likely to experience CDW than the low-risk group. On average, patients classified as high-risk experience more CDW events than the medium-risk group (HR=1.17, 95% CI: 0.96 to 1.43), but this was not statistically significant (p=0.11). When we looked at the model-derived risk scores (SunRiSe risk scores), for every unit standard deviation increase in the risk score at study entry, there was 0.36 (standard error=0.03) increase in EDSS at baseline (p<0.001). Over time, for every unit standard deviation increase in baseline risk, there was 0.02 increase (standard error = 0.07) in the rate of EDSS worsening (p=0.02).

#### Using a combination of MRI and clinical data at study entry to predict the risk at study entry

In the training set, when we added baseline EDSS, age and gender to MRI measures to predict 12-week CDW, the high-risk group had a hazard ratio of 1.75 (95% CI: 1.48 to 2.06) compared to the low-risk group (p<0.0001). This means that patients classified by the model as high-risk were approximately on average 75% more likely than the low-risk group to experience CDW during the follow-up. The medium-risk group had a greater hazard ratio of 1.19 (95% CI: 1.02 to 1.39) relative to the low-risk group, which was statistically significant (p<0.02). This means that the medium-risk group, on average, was approximately 19% more likely to experience CDW than the low-risk group. Similarly, the risk of developing CDW in the high-risk group compared with the medium-risk risk group was significantly higher (HR=1.46, 95%CI: 1.30 to 1.65, p< 0.0001). This means that patients classified as high-risk were 46% more likely to experience CDW than the medium-risk group.

When looking at the external test data set, the predicted high-risk group had a statistically significantly higher risk of EDSS progression in comparison with the low-risk group (HR=1.51, 95% CI: 1.18 to 1.92, p<0.0001) and the medium-risk group (HR=1.34, 95% CI: 1.11 to 1.62, p<0.01). This means that patients predicted to be in the high-risk group were 51% more likely than the low-risk group and 34% more likely than the medium-risk group and to experience CDW. More patients in the medium-risk groups than the low-risk group experienced EDSS progression (HR=1.12, 95% CI: 0.89 to 1.4), but this was not statistically significant (p=0.3). Figure 3 shows the number of patients in each group and the Kaplan Meier curves for the 12-week CDW. Figure 3 shows the Kaplan-Meier curves for the training and external validation data sets. Figure 2 shows the frequency of risk groups within clinical MS phenotypes.

**Figure 2.**
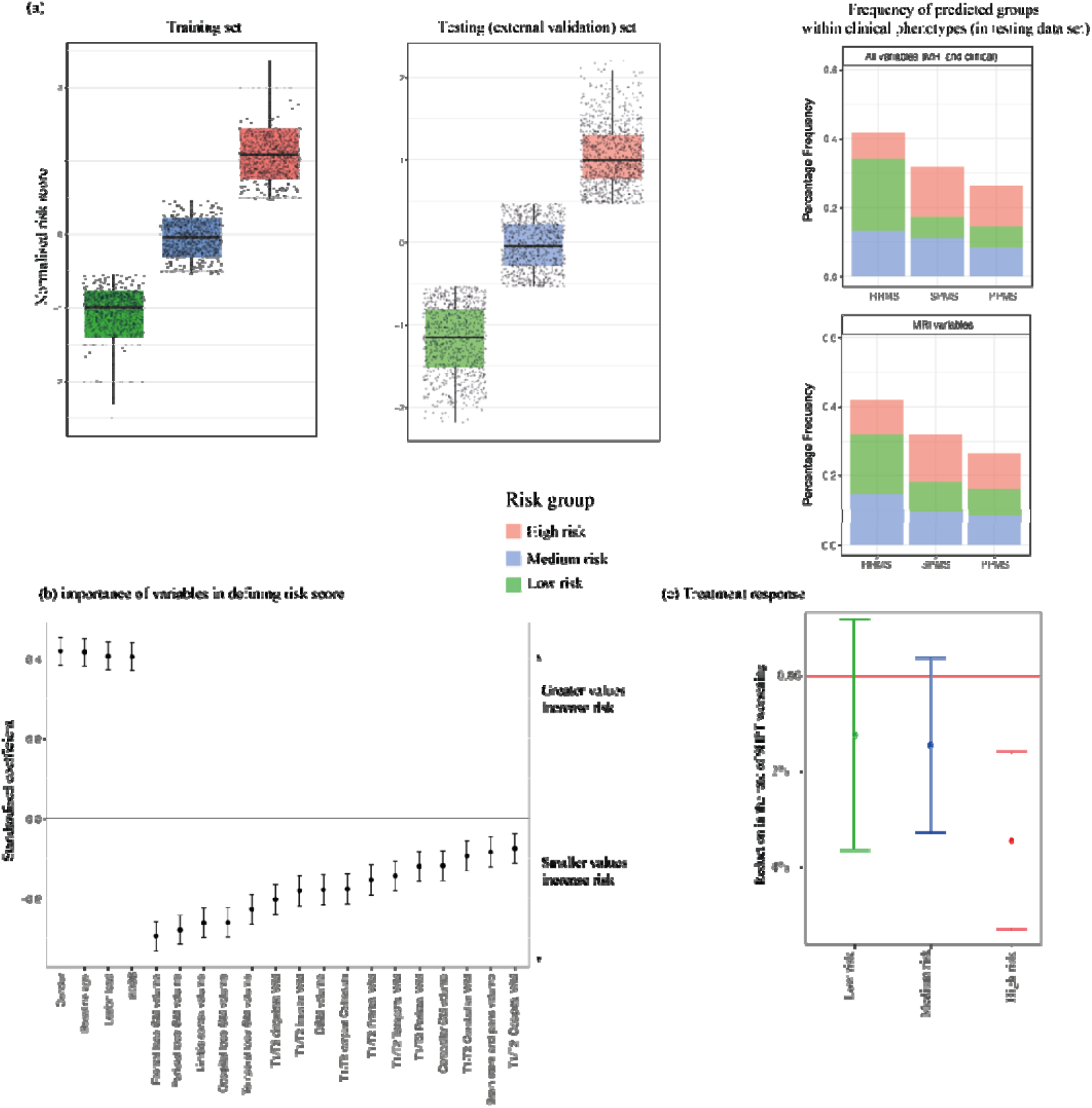
Population risk stratification and variable importance in defining the SunRiSe risk score. (a) shows the distribution of the three risk strata in the training and test dataset. The multi-level mixture models showed that three groups in the training set could best explain the data heterogeneity. We used the training set to normalise the risk scores (Z-scores) in the training and external validation data sets. The bar plots on the right-hand side show the frequency of risk groups within clinical phenotypes. When using clinical and MRI data, within the external validation set, a total of 50% of RRMS, 19% of SPMS, and 22% of PPMS patients were predicted to be at low risk. 31.8% of RRMS, 35% of SPMS, and 32.4% of PPMS were predicted to be at medium risk. 18.1% of RRMS, 45.9% of SPMS, 44.6% of PPMS were predicted to be at high-risk. When using MRI data, a total of 41.6% of RRMS, 25.9% of SPMS, and 27.8% of PPMS patients were predicted to be at low risk. 34.8% of RRMS, 30.5% of SPMS, and 32.9% of PPMS patients were predicted to be at medium risk. 23.4% of RRMS, 43.4% of SPMS, and 39.2% of PPMS were predicted to be at high risk. (b) Shows the variable importance analysis. The risk score was the outcome variable, and variables at study entry (horizontal axis) were predictors. Age, gender, lesion load, EDSS had a positive coefficient (greater values correspond to greater risk), while other variables had a negative coefficient (lower values correspond to greater risk score). Each unit increase in the variables in the horizontal axis corresponds with the standard deviation units (*Z*-score) of the risk score in the vertical axis. One standard deviation increase in EDSS score corresponds with a 0.4 standard deviation increase in the risk score while being a male corresponds to a 0.4 standard deviation increase in the risk score. (c) Shows the treatment response in ASCEND and ORATORIO trials. Patients in the high-risk group had a 4% annual reduction in the 9-Hole Peg Test worsening (absolute difference of 2.34 annual reduction compared to the placebo group, p=0.004). The error bars represent a 95% confidence interval. *Abbreviations:* GM, grey matter; DGM, deep grey matter; EDSS, expanded disability status scale.

**Figure 3.**
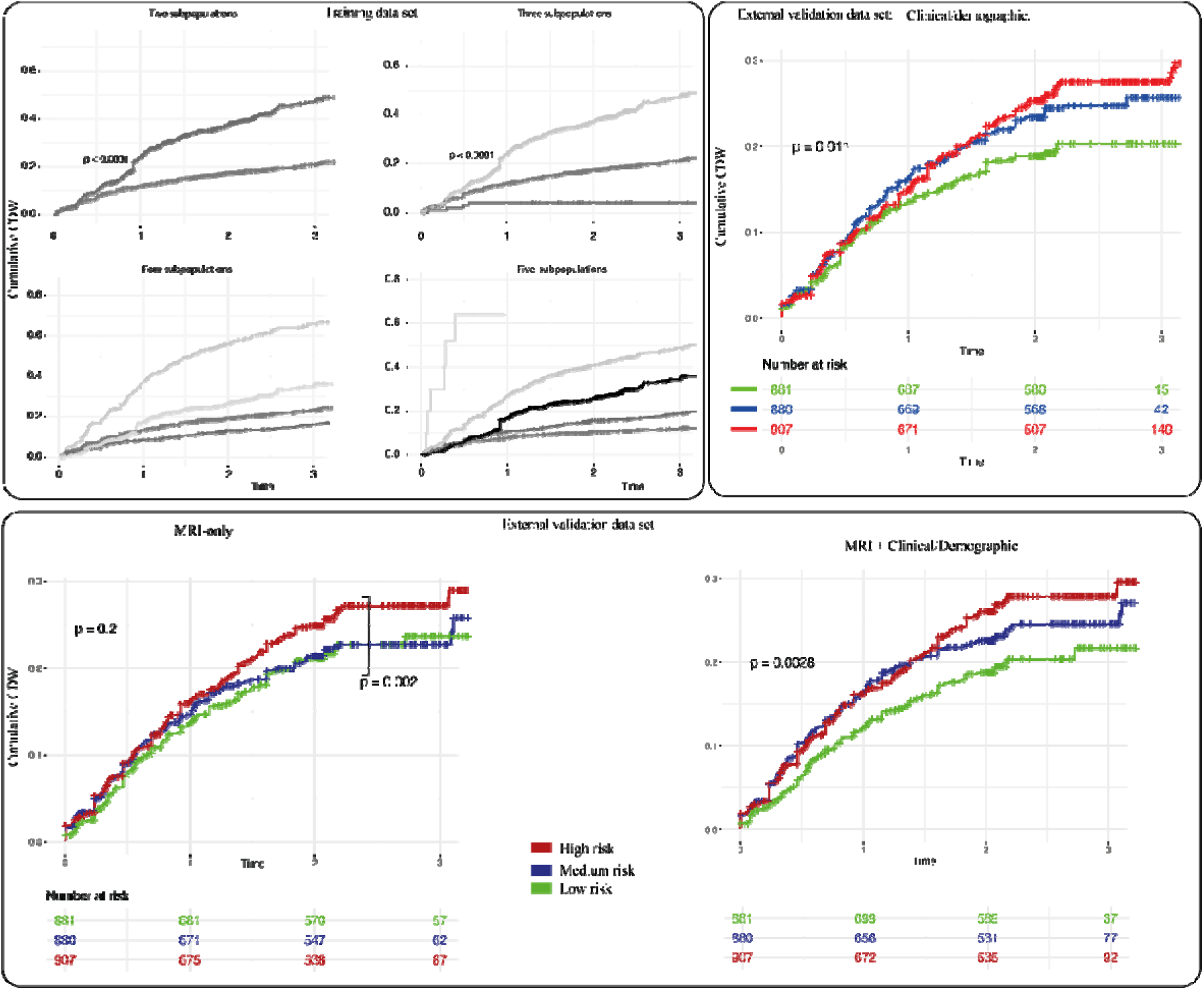
Stratifying subpopulations. The upper panel left panel shows the data-driven subpopulation inference using longitudinal data in the training data set. The three subpopulations could best explain the heterogeneity in EDSS worsening and time to CDW. We chose three as the optimal number of subgroup trajectories because by increasing the number of subgroups (4 and more), there were no statistically significant differences between all the pairs of subgroups. Additionally, by adding more than three subgroups, we did not find meaningful subgroups. We considered a subgroup to be “meaningful” when there were more than 100 patients and the difference in hazard ratios across subgroups was statistically significant (all comparisons in the training data set). The upper right panel shows the performance of the ensemble model in the external validation data set using *baseline clinical and demographic data (EDSS, age, gender, and disease duration)*. The lower left panel shows the model that was trained with MRI data only, and the right column shows the model that was trained with MRI, demographic and EDSS measures at baseline. The model that integrates both the clinical, demographic and MRI data has the best performance. The log-rank test p-value is shown alongside survival curves for each data set. In the lower left panel the p-value of the comparison of the hazard ratio between the high risk and low risk group is shown.

#### Importance of variables in defining the risk score

Variables measured at study entry that contributed to higher risk scores in order of importance were male gender, older age, higher lesion load, higher EDSS, lower GM volumes of the cortical lobes, lower T1/T2 in normal-appearing white matter in the cingulate and insular region, lower deep grey matter volume and lower T1/T2 of the corpus callosum (p=0.01 for all, see Figure 2b for the complete list). Our findings in Figure 2b and Table 2 show that increasing age, disease duration, baseline EDSS, lesion volume, male gender, smaller volumes of the cortex and deep grey matter, and lower T1/T2 in normal-appearing white matter contribute to and predict an increased risk of CDW.

**Table 2.** Subpopulation characteristics in the training and external validation data sets.

|  | ***Training and cross-validation***<br>*(n=5,483)* |  |  | ***External validation***<br>*(n=2,668)* |  |  |
| --- | --- | --- | --- | --- | --- | --- |
|  | **Risk group** |  |  | **Risk group** |  |  |
|  | **Low** | **Medium** | **High** | **Low** | **Medium** | **High** |
| Percentage of population<sup>1</sup> | 18 | 60 | 23 | 22 | 55 | 23 |
| Age (SD<sup>2</sup>) | 36.95<br>(9.59) | 42.59<br>(11.53) | 48.44<br>(9.26) | 37.38<br>(9.36) | 42.22<br>(9.47) | 47.20<br>(6.78) |
| Female (%) | 88% | 67% | 41% | 88% | 62% | 36% |
| EDSS<br>(Interquartile range)<sup>3</sup> | 2.5<br>(1.5 – 3.5) | 3.5<br>(2.0-6.0) | 4<br>(3.5-6.0) | 3<br>(2.0-4.0) | 4<br>(3-6) | 5.5<br>(4-6) |
| Disease duration (SD) | 4.7<br>(6.04) | 6.06<br>(6.36) | 8.12<br>(8.99) | 3.47<br>(4.58) | 6.01<br>(6.78) | 7.95<br>(6.93) |
| Lesion load in mL (SD)<sup>2</sup> | 13.29<br>(15) | 19.85<br>(21) | 25.08<br>(33) | 16.12<br>(16) | 21.90<br>(22) | 33.87<br>(34) |

<sup>1</sup> Percentage numbers are automatically rounded and therefore may not add up to 100%.
<sup>2</sup> Lesion load values are adjusted for age.
The model that was used to classify patients into risk groups used 21 measures (18 MRI variables, baseline EDSS, age and gender).
SD, standard deviation.

#### Disease activity: risk scores were not associated with relapses

When we used the model trained with MRI measures, there were no statistically significant differences in the annual relapse rate over time across the three risk groups in the external validation set (annual relapse rates were: low-risk=0.28, medium-risk=0.27, high-risk=0.25). When we used the model trained with MRI, age, gender and EDSS, there was a small but statistically significant difference across the three groups in the annual relapse rates. The low-risk group had the highest average (±standard error) annual relapse rate of 0.31 ± 0.02, which was statistically significantly different from the high-risk group with an average relapse rate of 0.22 ±0.03 (p=0.002). These two groups were not significantly different from the medium-risk group, which had an average annual relapse rate of 0.26 ± 0.03 (p=0.12).

#### Treatment response

In two clinical trials in the external validation data set (ASCEND and ORATORIO), patients in the high-risk group had a significant treatment response according to the rate of slowing of the 9-Hole Peg Test (p<0.001). The rate of 9-Hole Peg test performance in the high-risk group improved by 2.3 (±0.81) seconds per year on treatment, while it worsened by 3.13 seconds (±0.62) the high-risk patients on placebo (p=0.004). There were no differences in treatment response (difference between rate of 9-Hole Peg test performance between the placebo and treatment groups) between the medium and low risk groups. There were no differences in treatment response as measured by EDSS and the Timed Walk Test. Figure 2d shows the percentage change in the 9-Hole Peg test across the three groups and the treatment response.

#### Selecting patients predicted to be at high risk of disability worsening increased the statistical power compared to when all risk groups were sampled with equal weighting

##### Event-driven clinical trials

The average (±standard deviation) of the statistical power for a sample of 1,200 participants with three years of follow up, for an enriched trial of high-risk patients was 82% (±2.0) compared to 76% (±2.4) without patient enrichment. This corresponds to 266.71 ±14.6 CDW events for enriched trials enrolling high-risk patients compared to 224.31±13.4 events when patients with any level of risk are recruited (p<0.0001). This means that risk stratification and enrolment of high-risk patients reduced the duration of a phase 3, event-driven, clinical trial by an average of 4.5 months (±2.1 months).

#### Clinical trials with pre-defined sample sizes (conventional clinical trials)

In clinical trials with pre-defined sample sizes, the estimated sample of patients to achieve 80% power was 1,100, while without enrichment this was 1,300 participants to detect a 30% treatment effect on 12-week CDW. **Figure 4** shows the results of sample size calculations.

**Figure 4.**
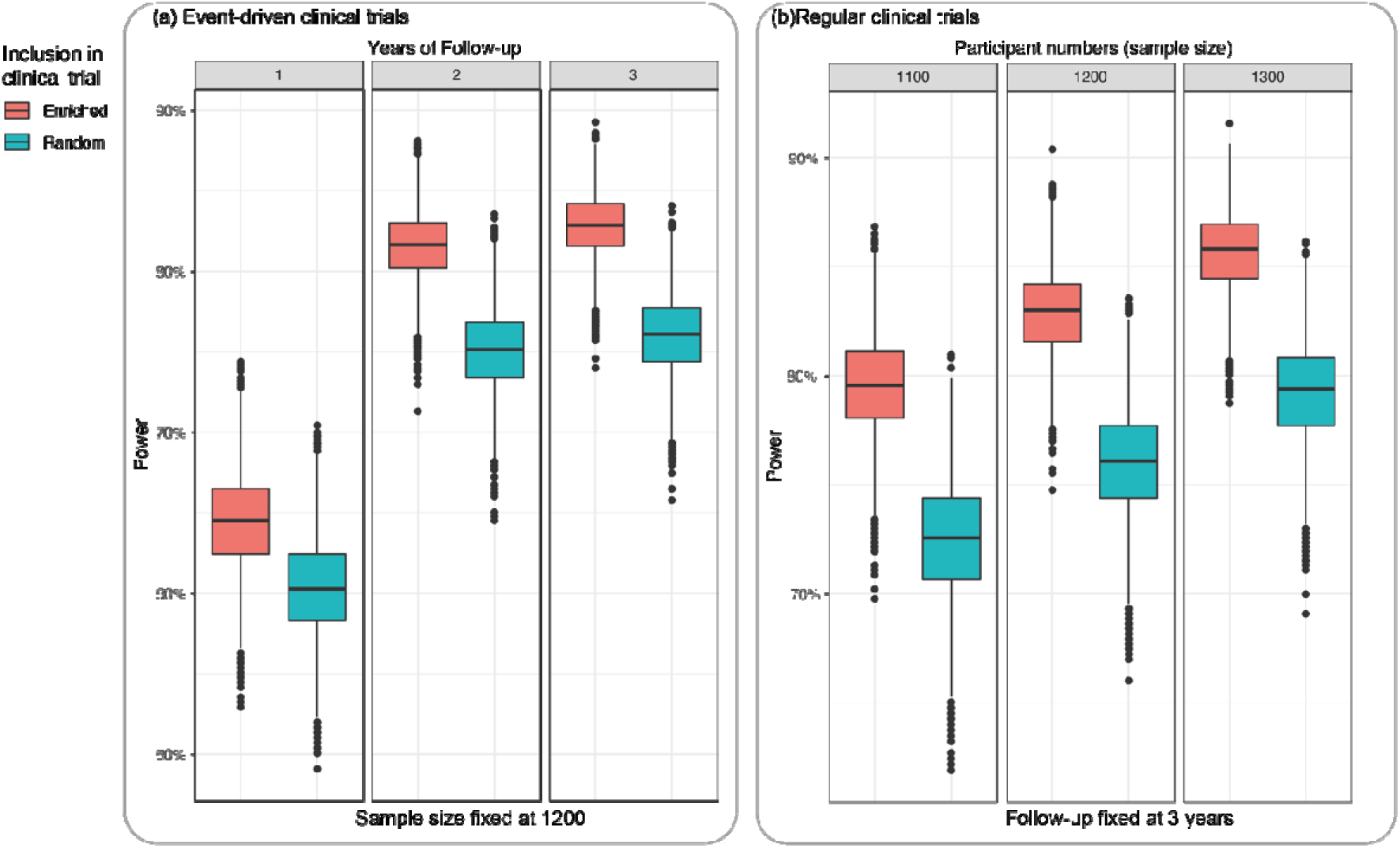
Statistical power of event driven clinical trials when patients at high risk of disability are selected. The figure shows that recruiting patients at high-risk of disability progression identified by the ensemble survival model significantly increase the statistical power of event driven (a) and regular (b) clinical trials. We used the external validation data set to sample 1,200 participants with replacement using baseline characteristics. To calculate the power, we used a hazard ratio of 0.7, and number of actual events based on the external validation data set.

## Discussion

We developed a risk score by combining baseline multi-modal imaging, demographic, and clinical data to predict disability worsening. Our risk scoring model generates values for individual patients. We defined risk groups based on individual risk scores in the training using both baseline and longitudinal data, and in validation*, using only baseline data,* we found that these groups had different risk of developing 12-week CDW when evaluated over time. When we applied our model to independent data we confirmed its generalisability and demonstrated its ability to increase statistical power and predict treatment response for clinical trials. Important variables at study entry that defined greater risk scores were male gender, increasing age, lesion load and EDSS, decreasing cortical volumes, decreasing deep grey matter volumes, and decreasing T1/T2 ratio in normal-appearing white matter. Our model can be used prospectively in clinical trials to screen and stratify patients across treatment arms.

Characteristics of subgroups between the training and external validation data sets were consistent. In these data sets, patients classified as high-risk were more likely to be men, older, more disabled at baseline, and have MS for longer. These are well-established risk factors for MS^7^ and are consistent with previous studies on EDSS, age, male gender^10, 14– 16, 44^, and radiological findings^8, 9, 53^. Most of these studies have used group-level statistical models to estimate the degree to which such factors contribute to long-term disability. The novelty of our work lies in our approach that exploits widely available quantitative neuroimaging data and provides individual-level risk stratification. Our study is unique in applying machine learning to large datasets with external validation and with the potential utility to increase statistical power in event-driven^48^ clinical trials that rely on the number of CDW events, thereby potentially reducing the duration of drug development. A downside is that about one-third of patients were predicted to be high-risk, and recruitment based on risk scores may increase the screening period of clinical trials.

We found that the loss of cortical and deep grey matter, a reduction in normal-appearing white matter T1/T2 ratio and lesion volume contribute to a higher risk score for disability worsening. We and others have previously shown that brain volume loss and lesion volume accumulation contribute to disability^37, 54^. T1/T2 ratio is a proxy for microstructural integrity, which may change because of several different underlying pathological processes, including demyelination^55, 56^. It is important to note that we adjusted MRI measures for demographic and clinical variables before entering them into our risk prediction models, to show the added value, rather than the MRI measures just acting as a proxy for demographic features. When looking at the relative importance of variables in defining the risk score extracted from the model, we found that in the order of importance, male gender, greater age, higher lesion load and higher EDSS were the most important variables. The volume of cortical lobes, T1/T2 ratio in the white matter surrounding the cingulate cortex, insular cortex, followed by deep grey matter, were the next important variables. Therefore, we can conclude that more extensive damage in normal-appearing white matter T1/T2 ratio, MRI-visible injury and grey matter volume loss contribute to our model-derived (SunRiSe) risk scores to different degrees independently of age, gender, and disability.

The risk scores were largely independent of relapse disease activity. There were no associations between annual relapse rate and risk scores. This can partly be related to our pre-processing steps and the outcome (CDW) we used to train our models. We excluded clinical assessments within (before or after) 30 days of a protocol-defined relapse. Recent studies have shown that more than 80% of the disability accumulation can be independent of relapses, even in relapsing-remitting MS^57^. Therefore, it is likely that the risk scores represent more of the neurodegenerative component and the underlying insidious progressive disability worsening across clinical phenotypes of MS, rather than relapse disease activity, and therefore there was no difference in response to anti-inflammatory treatments across the three risk strata. We did not find a significant reduction in the rate of disability worsening according to EDSS and Timed-Walk Test in treatment arms relative to the placebo arms in different risk groups. However, we found a small but statistically significant response to treatment in the high-risk group according to the change in the rate of 9-Hole Peg Test worsening in two clinical trials of SPMS and PPMS patients in our external validation data set. We believe that this relates to a more rapid decline in the hand function in patients who have less walking ability when entering trials (baseline EDSS for the high-risk group was 5.0), thus making it more likely for statistical models to detect a change. It has been previously reported that hand function may be more sensitive in assessing treatment response in more disabled populations whose walking function may not change^24^. Patients classified as high-risk had higher disease burden (according to higher lesion load and lower brain volumes) at study entry but a less active disease according to relapse rates over time. This means that these patients had a higher disease burden to start with but a less active disease and were more likely to experience disability worsening. In our previous work, using the same cohort of patients, we found a significant treatment response when patients were grouped based on MRI-derived patterns ^17^. In our previous work, we predicted CDW by using the Subtype and Stage Inference or SuStaIn algorithm on MRI to classify patients into data-driven MRI subtypes. We found that an MRI-derived group coined lesion-led had a higher disease activity, faster disability progression, younger age, and greater treatment response^17^. SunRiSe risk scores introduced here for the first time could predict CDW more strongly than our previous model, though not treatment response. The largest hazard ratio across MRI-based subtypes was 1.32 for the SuStaIn model, while for the SunRise model, this was 1.51. This is because SunRiSe is a supervised survival model trained to predict time to CDW directly. The factors that contribute to higher risk of disability act in opposite directions in defining treatment response to anti-inflammatories. For example, older age contributes to higher disability but reduces treatment response, and a more active inflammatory disease (higher/new T2 lesion) increases disability accumulation but increases the likelihood of responding to anti-inflammatory therapies. We believe this is a reason why the SunRiSe model could not predict treatment response as well as SuStaIn, while outperforming SuStaIn in predicting future disability. Overall, our findings across these two studies mean that the risk-groups based on the SunRiSe ensemble model and our previous SuStaIn model distinguish patients with different characteristics, thereby they have the potential to complement each other in future works.

We used three different types of survival machine learning models to address potential pitfalls of one with another and enable symbiosis in our ensemble model to achieve superior performance compared to using any one model on its own (Figure 3 and Supplemental Table 1). Deep learning models are known to be sensitive to centre effects and changes in data distributions. To alleviate this issue, we used two other models (mixture models and gradient boosting survival trees) multi-centre datasets (details of centre effects explained elsewhere^17^). When we applied our model to independent, external cohorts, our results were reproduced, which underpins the robustness of our model to changes in scanner and centre.

Models developed only with MRI measures were able to distinguish across two (high risk vs low risk) of the three risk groups as shown in **Figure 3**. Adding EDSS along with age and gender increased the performance of our model (maximum hazard ratio in the MRI-only model was 1.69 while it was 1.75 in the model that included both MRI and clinical data) and could distinguish across the three groups. Our MRI-extracted variables were adjusted for age, disease duration and EDSS, and therefore present independent contributions from the demographic values. A recent study showed that age at onset and EDSS are important predictors of ‘aggressive’ multiple sclerosis ^14^. Several previous studies have shown that higher EDSS and age are predictors of a worse prognosis. However, it remained unclear whether quantitative MRI measures which can be extracted from widely available MRI sequences may add value to clinical variables.

Although SunRise risk scores are predicted for individuals, our models are still not suitable for robust prediction of disability worsening at an individual level, but still moves us from a group-wise approach towards a actionable subgroup-based risk prediction. We trained our model to explain disease worsening according to EDSS (we observed similar results in the 9-Hole Peg Test and Timed-Walk Test, results not shown) which has well recognised limitations 58 which may in part explain the still limited predictive power at an individual level. Future research on emerging measures of disability will provide outcomes with lower variability and greater neural network specificity, and so may enable more personalised predictions. However, we believe that using EDSS is also a strength our study, because of its wide acceptance by regulators across the world which in turn enables our models to have a wider clinical impact in the future.

In conclusion, we developed a risk stratification model and predicted short-term disability worsening using a combination of neuroimaging and clinical measures. Our model generalised to unseen, independent cohorts of patients in phase two and phase three clinical trials and can be used prospectively to stratify the risk of disability worsening.

## Data Availability

The data sets are controlled by pharmaceutical companies. Requests to access data can be forwarded to data controllers listed in our previous publication17. Processed CSV files can be from the corresponding author by any qualified investigator for reproducing the results of this study. 

## Acknowledgement

This investigation was supported (in part) by (an) award(s) from the International Progressive MS Alliance, award reference number PA-1412-02420. We are grateful to all the IPMSA investigators who have contributed clinical trial data to this study as part of EPITOME: Enhancing Power of Intervention Trials Through Optimised MRI Endpoints network (see the list of investigators in the appendix). This study was also supported by the National Institute for Health Research University College London Hospitals Biomedical Research Centre. O.C. is a National Institute of Health Research (NIHR) Professor (grant code: RP-2017-08-ST2-004). We thank Rozie Arnaoutellis, Istvan Morocz, and Caramanos Zografos for coordinating and organising this study. We thank Jonathan Steel and Jonathan Stutters for their IT support during this work. D.C.A. has received funding for this work from Engineering and Physical Sciences Research Council Grants M020533, M006093, and J020990. This project has received funding from the European Union’s Horizon 2020 Research and Innovation Programme under Grant Agreements 666992. MS-SMART is an investigator-led project sponsored by University College London (UCL). PAW was supported by an MRC Fellowship (MR/T027770/1). NPO is a UKRI Future Leaders Fellow (MR/S03546X/1). N.P.O, O.C, D.C, F.B. and D.C.A acknowledge funding from the National Institute for Health Research University College London Hospitals Biomedical Research Centre. PAW was supported by an MRC Fellowship (MR/T027770/1). NPO is a UKRI Future Leaders Fellow (MR/S03546X/1). MS-SMART project (reference 11/30/11) was funded by the Efficacy and Mechanism Evaluation (EME) Programme, a Medical Research Council (MRC) and National Institute for Health Research (NIHR) partnership. The views expressed in this publication are those of the author(s) and not necessarily those of the MRC, NIHR, or the Department of Health and Social Care. A.J.T. is a National Institute of Health Research (NIHR) Emeritus Senior Investigator.

## Supplemental methods

We discuss the methodological details of the three models that contribute to a final risk score (SunRiSe risk) in this section. In all these models EDSS, a well-established outcome accepted by regulators and used in clinical trials, was the clinical variable of interest. The three models that constituted the ensemble model included two supervised models (1 and 3) and one unsupervised classifier (model 2) as below:

### 1) Deep neural network model: DeepHit

We used the original implementation^3^ of DeepHit architecture. DeepHit learns the joint distribution of CDW and the time to CDW while taking into account the right censoring of data for those who completed (or were dropped out of) the study without experiencing CDW. DeepHit makes no assumptions about the form of underlying data distributions and that of the relationship between covariates and the hazard rate. This enables learning non-linear relationships between covariates and hazard rates, thus outperforming some of other deep neural network-based architectures such as DeepSurv that rely on Cox-regression model assumptions.^58^ We hypothesised that the ability of learning non-linear relationships will provide a boost in predicting the disability because of the well-known bimodal distribution of EDSS.^59^

### 2) Trajectory-based classification and risk scoring with longitudinal multilevel mixture models

Mixture models are an unsupervised algorithm that we used to classify the trajectory of EDSS worsening. Here we aimed to separate subgroups of patients whose rate of disability worsening measured by EDSS differed. The justification for using clustering of subgroups, as opposed to individual-level prediction, was the well-known limitations of EDSS in having a bimodal distribution and relatively high variability and lower sensitivity to subtle disease worsening. This enabled us to use EDSS as the outcome and choose the optimal number of subgroups in a data-driven way to maximise generalisability. During the training, we applied longitudinal mixture models to the EDSS data to explore the optimal number of subpopulations with similar worsening rates over time^45^. We performed leave-one-out cross-validation to select the optimal number of subgroups that could best explain different trajectories of EDSS worsening (trajectory-based classification). We iterated the cross-validation for ten times, each time fitting the model on 9 folds and applying it on the remaining, held-out fold. We chose the optimal number of subgroups as the one providing statistically significant hazard ratios across all data-driven subgroups in the training data set and cross-validation folds. After choosing the optimal number of subpopulations, in the second step, we trained a gradient boosting classifier based on the cross-sectional characteristics of the members of each subtype at study entry (baseline). For each subgroup, we used a Cox model to generate risk scores for individuals. In the external validation, we only used the trained cross-sectional classifier to define the most likely subgroup membership and weighted the Cox model predictions according to the membership probability. Ranking the outputs of the Cox model allowed to generate individual risk scores.

### 3) Gradient boosting survival trees

Gradient boosting survival model is an ensemble of sequential weak decision trees that provide boosted performance and has extensively been used in classification, regression and survival models.^47^ They are similar to random forest models (except that they use sequential trees), with robust performance in defining variable importance when categorical (e.g., gender) and ordinal (e.g., EDSS) variables are used.

#### Consensus risk score and subgroup stratification

Each of the above three models provides a risk score of developing 12-week CDW. To combine the outputs of the three predictive models into one risk score, we used a linear model (meta-model) that received the risk scores from individual models and provided a consensus risk score. We grouped patients based on the consensus risk score into three groups of high, medium and low risk score based on the consensus risk score tertiles that we defined in the training data set. To prevent bias and inflating false positives, we did not use the external validation in defining the threshold.

## Supplemental results

### Ten-fold cross-validation

The two subgroup and three subgroup models showed statistically significant differences in all cross-validation folds across the subgroups. Therefore, we chose the three groups as the optimal model to best classify the differing subgroup trajectories. The results of mixture models with up to 5 subpopulations applied to all 10-folds in the training set is shown in Figure 3 (upper panel).

---

**Supplemental Table 1.** Results for individual models and the ensemble (SunRiSe) model.

|  | High-risk vs low risk | p-value | High-risk vs medium-risk | p-value |
| --- | --- | --- | --- | --- |
| Stratified<br>predictions<br>(mixture<br>model) | 1.41<br>(1.10-1.57) | P=0.005 | 1.32<br>(1.10-1.57) | P=0.002 |
| DeepHit | 1.54<br>(1.25 -1.91) | P<0.001 | 1.40<br>(1.13-1.71) | P=0.001 |
| GBS* | 1.19<br>(0.98-1.45) | P=0.08 | 1.03<br>(0.84-1.27) | P=0.7 |
| Ensemble<br>model | 1.75<br>(1.48-2.06) | P<0.0001 | 1.46<br>(1.30-1.65) | P<0.0001 |

\*Gradient boosting survival trees

## Collaborators and investigators of the International Progressive MS Alliance (PMSA)

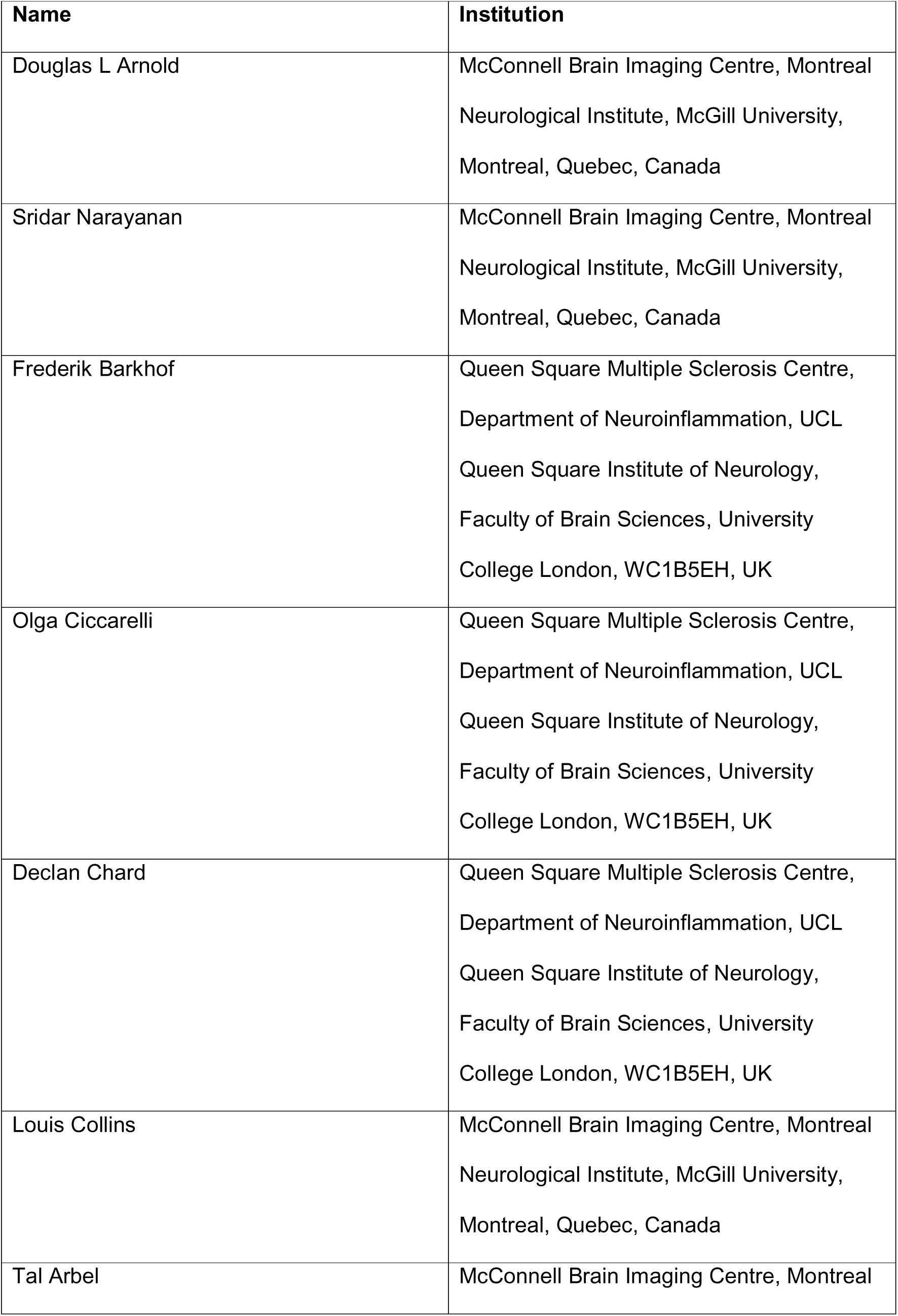
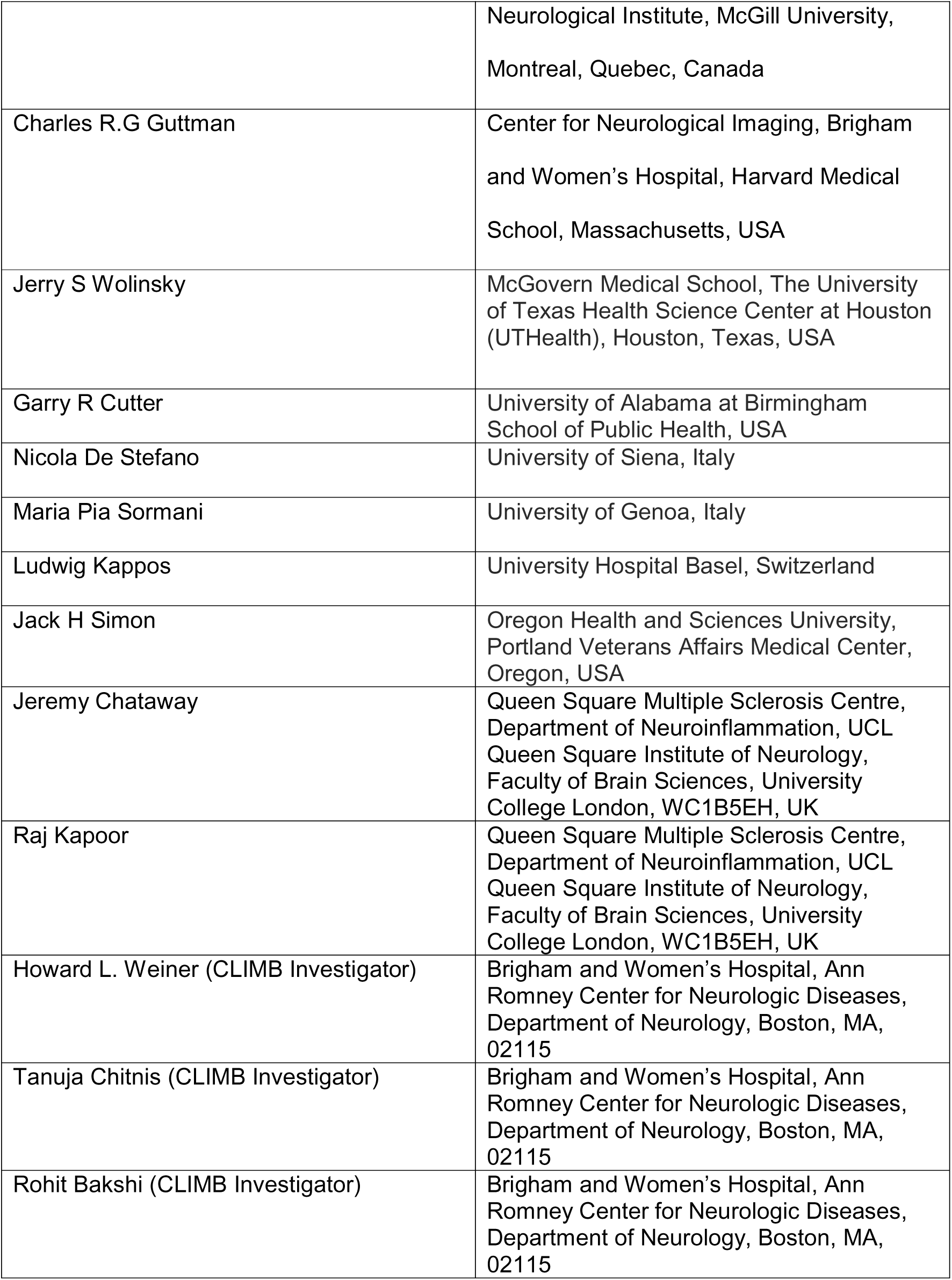

---

**Supplemental Figure 1.**
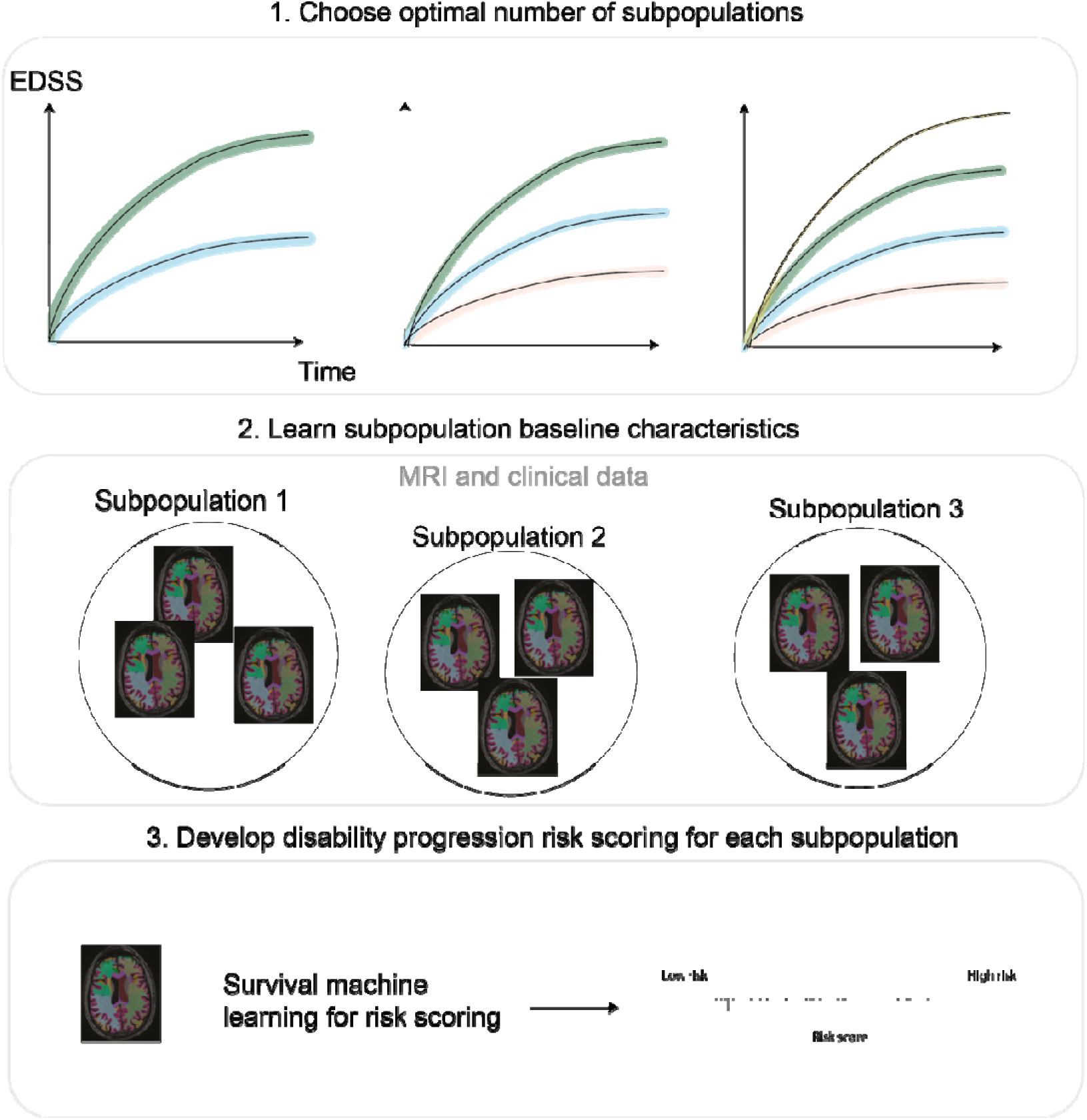
**Flowchart of the** model-based stratification.

**Supplemental Figure 2.**
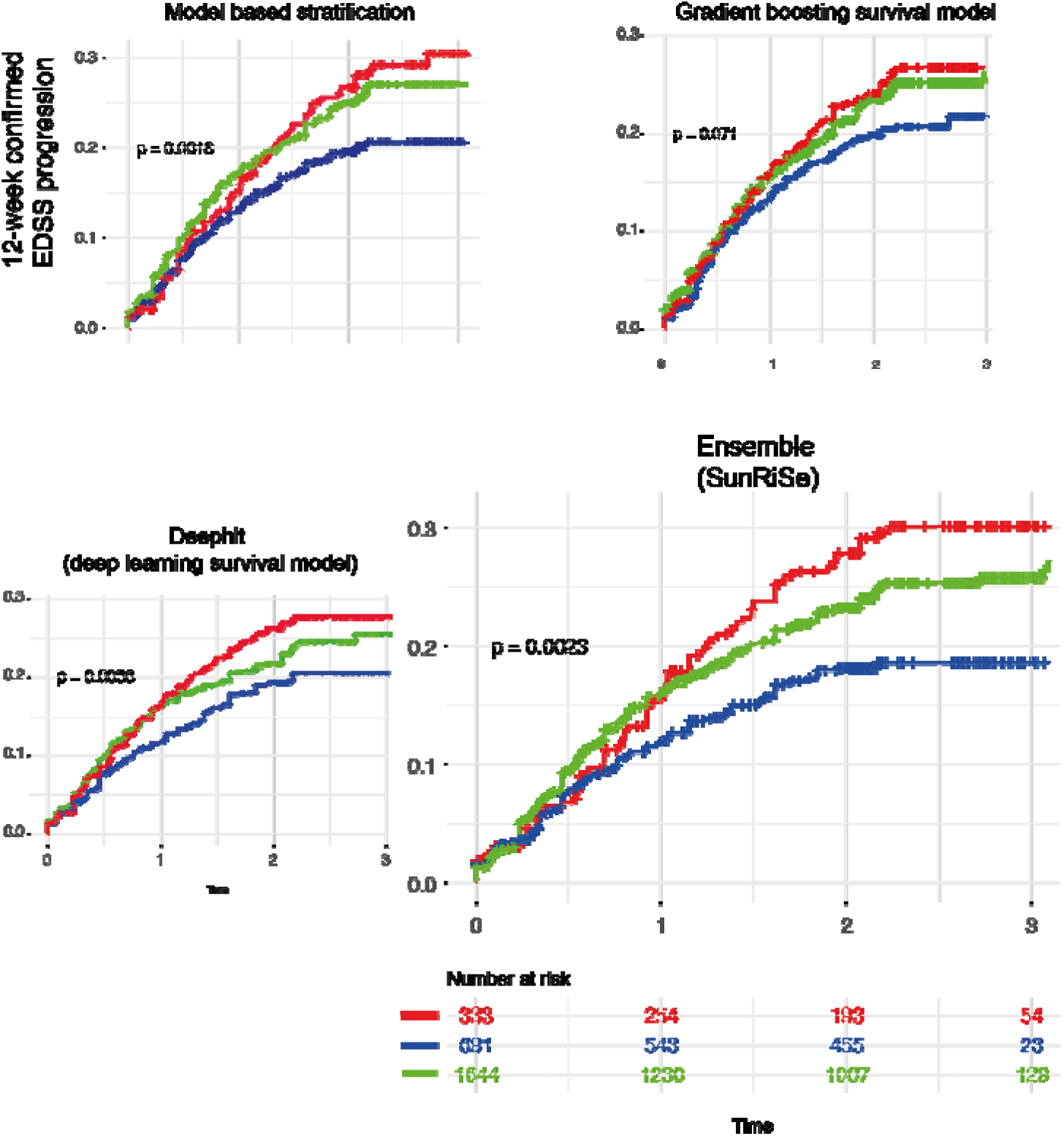
Predicting disability in external validation data set with individual models.

